# The training specificity versus structural adaptation paradox: Differential effects of isokinetic concentric and eccentric resistance training on muscle architecture and function in young men

**DOI:** 10.1101/2025.03.31.25324923

**Authors:** João Pedro Nunes, Kazunori Nosaka, Anthony J. Blazevich

**Affiliations:** School of Medical and Health Sciences, Edith Cowan University. Joondalup, Australia

**Author notes:** Corresponding author: João Pedro Nunes. School of Medical and Health Sciences, Edith Cowan University. Joondalup, Australia.

**Keywords:** Strength training, Shortening, Lengthening, Sarcomerogenesis, Muscle gear, Muscle geometry, Rate of force development, Stretch-shortening cycle

## Abstract

It is unclear whether muscle functional adaptations to concentric (CON-RT) and eccentric (ECC-RT) resistance training are most specific to their exercise characteristics or the structural adaptations they evoke. In this study, the effects of effort- and volume-matched CON-RT and ECC-RT on regional hypertrophy, muscle architecture, and function were compared, and associations between the outcomes were explored. Twelve trained young men (25.5±3.6y) completed 18 isokinetic ankle-dorsiflexion exercise sessions over 6 weeks: CON-RT in one leg and ECC-RT in the other (2-4 sets, 6-10 maximal repetitions, 10°/s). Tibialis anterior size and architecture (ultrasound imaging) and maximum voluntary dorsiflexion function (isokinetic dynamometry) were assessed. Muscle thickness increased similarly between conditions and across proximal-distal regions (8%), pennation angle increased more in CON-RT (8%) than ECC-RT (4%), and fascicle length increased only after ECC-RT (7%). Functional adaptations were more closely associated with specific structural adaptations than with contraction mode, velocity, or angle. Isometric torque increased similarly in both conditions overall (8%) but CON-RT improved only at shorter muscle lengths and shifted the peak-torque angle leftward, whereas ECC-RT improved at both shorter and longer lengths and broadened the torque-angle plateau, which was associated with fascicle length increases. ECC-RT produced greater increases in both eccentric (13%) and concentric torques (17%) than CON-RT (3%, 9%, respectively), and changes were similar across velocities – contrary to the training specificity theory. Changes in pennation angle were associated with dynamic strength changes. These findings suggest that muscle function adapts to the structural changes induced by training, regardless of the training scheme used.

## 1 INTRODUCTION

A skeletal muscle’s architecture is largely characterized by its fascicle lengths (FL) and pennation angle (PA), defined as the lengths of the muscle fiber bundles and their angle relative to the muscle’s line of action, respectively [1,2]. Together with size descriptors such as muscle thickness (MT) or cross-sectional area, these architectural features determine a muscle’s length- and velocity-dependent force production characteristics [1,2]. Specifically, longer fascicles allow for increased maximum shortening velocity and greater force generation across a wider excursion range [1,3], whilst greater pennation enables a higher force production per volume [2] and enhanced rates of force development [4].

From this perspective, it follows that exercise interventions modifying these architectural features should predictably affect muscle function [5]. However, training-induced architectural adaptations present a paradox. While the training-specificity principle suggests that concentric movement velocity should be best improved by fast concentric training, increased FL, which is fundamental to muscle shortening velocity due to the increased number of sarcomeres in series, is commonly suggested to be best achieved by eccentric training, even at slow speeds [6]. Similarly, although eccentric training should best improve peak force production due to the higher force capacity of eccentric contractions, the requisite increases in physiological cross-sectional area are best achieved through increasing PA, which seems to predominate after concentric training [6]. Hence, a “training specificity” versus “structural adaptation specificity” paradox emerges.

Research exploring these relationships has produced mixed results. Blazevich et al. [7] found similar increases in vastus lateralis PA, FL, and muscle volume between concentric and eccentric isokinetic knee extension training programs, contradicting common hypotheses, yet found that changes in FL were positively associated with increased torque at longer muscle lengths, as expected [3]. In contrast, Quinlan et al. [8] observed greater vastus lateralis PA increases after concentric isoinertial leg-press training and greater FL increases after eccentric isoinertial leg-press training, as often hypothesized, yet found that FL gains were associated with torque improvements at shorter, rather than longer, muscle lengths. Moreover, others have reported similar functional adaptations between training modes despite differential architectural adaptations being triggered by concentric versus eccentric training across a range of muscles [9–11]. Therefore, it remains unclear whether training specificity or adaptation specificity predominates after voluntary exercise training in humans, which has important implications for the design of training programs to evoke specific functional outcomes.

One important consideration is that previous studies have typically analyzed architectural parameters in a single region of one muscle within a larger synergistic group and then explored their relation to whole muscle group function, despite evidence that architectural parameters vary within and between synergistic muscles at rest [12], during contraction [13], and in response to training [14]. A more comprehensive approach examining regional changes across multiple synergistic muscles, or within single muscles that contribute almost exclusively to torque production at a joint of interest, might better elucidate the *in vivo* architecture‒function relationship [15]. Additionally, many studies have primarily focused on changes in maximum strength, neglecting other functional parameters theoretically influenced by architectural adaptations, such as torque‒angle‒velocity characteristics and rate of contractile force development, which require further investigation.

Another key limitation of previous studies is the lack of standardization across training parameters. Studies have either compared maximal-effort concentric training to submaximal eccentric training matched for volume, or used equal relative intensities but a higher volume of eccentric training [7–11,16]. Given that contraction effort strongly influences force-related adaptations [17,18] and training volume is critical for muscle morphological changes [19], the lack of standardized training intensity and volume complicates result interpretation. Historically, only a few studies have equated volume between conditions whilst maintaining maximal effort contractions, but they did not assess changes in muscle architecture [20,21].

Given these considerations, the present study compared the effects of intensity of effort- and volume-matched concentric (CON-RT) and eccentric isokinetic resistance training (ECC-RT) programs on regional muscle architecture and a comprehensive range of muscle functional parameters assessed during isometric, concentric, and eccentric contractions. Associations between changes in architecture and changes in function were then explored. The tibialis anterior was selected for the study because of its predominant contribution to dorsiflexion force [22]. A within-subject study design to compare one leg with CON-RT and the other leg with ECC-RT was employed to minimize confounding factors such as diet, recovery, and exercise tolerance. Trained individuals were selected for their familiarity with high-intensity training, but they had no prior tibialis-specific training experience to avoid adaptation bias. It was hypothesized that strength adaptations would follow the specificity of structural adaptations.

## 2 MATERIALS AND METHODS

### 2.1 Study design

Anamnesis questionnaires and familiarization of the exercise and testing procedures were completed on a first laboratory visit, followed 7-10 days later by pre-training testing of tibialis anterior architecture and function. The training program commenced 3-5 days hence, lasting 6 weeks (18 sessions), which have been shown to be sufficient to induce morphological and functional adaptations [10,20,21], and post-training testing was conducted 5-7 days after the final training session. Participants’ legs were randomly allocated to CON-RT or ECC-RT conditions, balanced for dorsiflexion strength levels. To achieve this, participants were tested for their maximum isometric, concentric, and eccentric dorsiflexion strength levels before the training commenced, and legs were classified as ‘stronger’ or ‘weaker’ based on the total strength index calculated as the sum of the three strength scores [23]. Each participant was assigned a random number in an Excel list, which determined whether the stronger leg would follow the CON-RT or ECC-RT; therefore, half of the participants completed CON-RT with their stronger leg and ECC-RT with the weaker leg, and vice-versa.

Tibialis anterior architecture was assessed using ultrasonography followed by dorsiflexion torque‒ angle‒velocity parameters using isokinetic dynamometry, before and after training at the same time of day by the same investigator. Participants were instructed to refrain from intense lower-limb exercise for 48 h before testing sessions and to maintain their physical activity and diet habits throughout the study. A diet log was completed for the 3 days before pre-training testing to allow for replication after the training period. Pre- and post-training diet logs were used to calculate the average dietary intake using a diet tracking app (MyNetDiary Inc.). The study followed the Declaration of Helsinki, was approved by the University Ethics Committee (2023-04354-NUNES), and commenced after participants signed an informed consent form.

### 2.2 Participants

The required sample size was estimated *a priori*using G-Power software (v.3.1.9.6) considering an F test with α value of 0.05, power of 0.95, and large (0.80) Cohen d effect size, for two groups and two measurement times. A total of 24 subjects was required, but considering 25% potential dropouts and the within-subject design used, in which each leg is considered a sample unit, 15 subjects (n = 30 legs) was considered the target recruitment number. Recruitment was completed using social media and home delivery of flyers in the university area. Interested individuals completed detailed health history and physical activity questionnaires, and were subsequently admitted if they met the following inclusion criteria: adult 18-35 years old, no history of injury or musculoskeletal disorder at the ankle or related musculature, no regular use of non-steroidal anti-inflammatory drugs, no medical contraindication to high-intensity exercise, and involved in consistent resistance training practice for >1 year without specifically training the dorsiflexors. Study participation was not restricted by sex; however, only men volunteered to participate, and the required sample size was completed. Of 21 initial volunteers, 16 met the inclusion criteria and completed the familiarization session. Three decided not to participate, so 13 participants commenced the training. Having a training attendance of <80% or missing two sessions in a row was defined as an exclusion criterion, which eliminated one further participant. Therefore, 12 young men (24 legs) completed the study and were included in the final analyses (age = 25.5 ± 3.6 years; body mass = 79.3 ± 14.3 kg; stature = 179.2 ± 6.7 cm; BMI = 24.6 ± 3.7 kg/m²; habitual physical exercise frequency: 5 ± 2 d/wk).

### 2.3 Muscle architecture and size

Participants laid supine on a plinth with ankles at 0° (perpendicular to the floor) fixed in a metallic platform and rested for 10-15 min before measurements commenced. The proximal (anterolateral surface of the lateral tibial condyle) and distal (mid-shin) superficial tibialis anterior endpoints were identified using a 50-mm linear ultrasound probe (Prosound F75; probe model: UST-567 [5-15 MHz]; Hitachi Aloka, Japan) and these points marked on the skin with a dermatographic pen. Medial and lateral muscle boundaries were identified via ultrasound imaging along the whole muscle, a muscle mid-sagittal line was marked across the skin muscle-end marks, and transverse lines were marked at 25%, 50%, and 75% of muscle length. After water-soluble gel (Aquasonic 100; Parker Laboratories, USA) was generously applied between the probe and the skin to avoid compressing the skin and deforming the muscle, ultrasound images (gain = 80 dB; brightness dynamic range = 70; gamma curve = linear; depth = 35-45 mm, individually adjusted) were obtained at these three regions with the probe placed longitudinally to the tissue interface.

The probe was subsequently manipulated (tilt, rotation, yaw axes) to ensure optimal fascicle and aponeurosis visualization, and image acquisition was repeated until deemed of good quality. Images of the three regions were stored and later collaged using imaging software (PhotoFiltre Studio; France) with care to pair aponeuroses, fascicles, and anatomical landmarks. Subsequently, muscle architecture parameters were analyzed at proximal (33%) and distal (66%) regions of the muscle using ImageJ software (NIH, USA). FL was measured using the ‘segmented length’ tool on ImageJ as the distance between the ends of the most continuously visible hyperechoic line crossing the center of the region of interest and considering the fascicle’s non-linear path.

MT was calculated as the average distance between the superficial and central aponeuroses of the outlined fascicle. PA was calculated as the average angles formed between the superficial and central aponeuroses and the straight line connecting fascicle ends. Figure 1 shows an example of the ultrasonography images.

**Figure 1.**
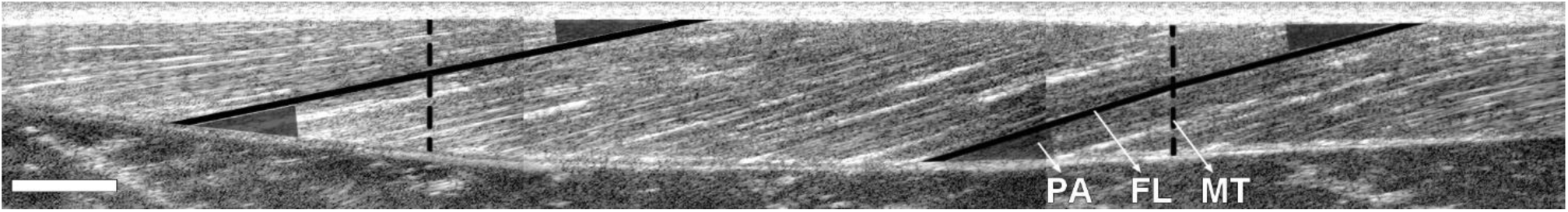
Example image collaged from the three ultrasound images obtained from the superficial tibialis anterior (distal-proximal). Shaded triangles = pennation angle (PA); solid lines = fascicle length (FL); dashed lines = muscle thickness (MT). White box scales for 1cm.

### 2.4 Muscle function

Participants were asked to perform a light warm-up consisting of two 10-s ankle dorsiflexor and plantarflexor stretches and 20 active ankle circumductions before being seated on the adjustable chair of the isokinetic dynamometer (Biodex System 4; USA), with hip at 85° (anatomical position = 0°) and knee fully extended. Inelastic straps were placed across the thigh and dorsum of the shoeless foot (above the toes), tightly strapped to the dynamometer footplate with the lateral malleolus aligned to the dynamometer’s axis of rotation. The dynamometer settings were recorded individually during the pre-training testing and kept constant during post-training testing. Participants performed six progressive 3-s, submaximal, isometric dorsiflexion contractions (ankle at 0°; foot sole perpendicular to the floor) as a further warm-up.

For testing, participants performed a series of one-repetition maximal voluntary isometric contractions with the ankle at -5° (i.e., 5° of dorsiflexion), 0°, 20°, 40°, and 45° plantarflexion, and in eccentric and concentric modes at angular velocities of 10°/s, 45°/s, and 90°/s. For all tests, participants were instructed to contract as fast and forcefully as possible. For the isometric tests, participants held contractions for 2-3 s, with torque plateau typically being achieved within 1 s. For the dynamic tests, participants completed an eccentric-concentric contraction pair from -5° to 45° plantarflexion and back, preceded by a 2-s maximal-effort isometric contraction at -5°, at each velocity. They then performed a concentric-eccentric contraction pair from 45° to -5° and back, preceded by a 2-s isometric contraction at 45°, at each velocity. This allowed the evaluation of concentric and eccentric functions preceded by an isometric contraction as well as by the opposite muscle action (i.e., concentric preceding eccentric, and vice versa). Verbal encouragement and visual feedback (large TV screen in front of the participants with torque-angle data shown in real time) were provided to promote maximal effort. Rest intervals between all tests were 1-2 min. The leg to be tested was first randomly selected, and participants were afforded ∼20 min of rest after the completion of all attempts on one leg before tests were performed using the other leg. Dynamometer torque, angle, velocity, and time data were captured at a 1000-Hz analog-digital sample frequency using PowerLab hardware connected to a computer using LabChart software (Powerlab System; ADInstruments, Australia).

From the isometric test data, the following parameters were assessed: (i) isometric peak torque at the five joint angles; (ii) angle of peak torque; (iii) width of the plateau region of the torque-angle relationship, defined as the angular distance over which torque remained within 2.5% of the peak torque value; and (iv) rates of torque development (RTDs) computed from torque onset to 75 ms and 250 ms, which were subsequently normalized to the peak torque (rRTD), with the last three variables computed using data filtered using 3^rd^-order polynomial functions.

From the concentric test data at the three velocities and two contraction histories, the following were assessed: (i) concentric peak torque at 20° dorsiflexion (see details below), (ii) concentric work calculated as the product of torque and angular displacement, and (iii) maximum concentric angular velocity, estimated from torque‒velocity linear regression on peak torques, all after filtering using 3^rd^-order polynomial functions. Concentric maximal voluntary function was assessed through both peak torque and work to capture different aspects of force production, and, since peak torque in pre-activated concentric tests typically occurs immediately after the preceding isometric or eccentric contraction due to force depression phenomena [24], peak torque was measured at a standardized joint angle of 20° dorsiflexion to better capture the concentric force production potential of the muscles and to allow for estimation of maximum shortening velocity from torque captured at a consistent joint angle.

From the eccentric test data, the following were assessed: (i) eccentric peak torque at the three velocities and contraction histories, and (ii), eccentric peak torque normalized to isometric torque, to identify whether eccentric strength changes were driven by changes in the preceding isometric torque or by changes in torque during active lengthening itself. This ‘net eccentric torque’ was calculated as the change (in Nm and %) in peak torque from the isometric phase to the eccentric phase at each velocity, and the average value across velocities was then computed.

For all contractions, active torque was calculated by subtracting the resting torque values, recorded across the full range of motion and filtered with 3^rd^-order polynomial functions, from the raw ankle joint torque values. Torque‒angle‒velocity‒time records obtained during the tests are illustrated in the supplementary material.

### 2.5 Training program

A supervised training program was completed thrice weekly (Mondays, Wednesdays, and Fridays) for 6 weeks. Participants were trained on the isokinetic dynamometer using the same chair settings and warm-up protocol as testing sessions. Maximal-effort concentric or eccentric contractions were performed through a 0° to 40° plantarflexion range of motion at an angular velocity of 10°/s. After each contraction, participants relaxed the foot, and the platform was returned (passively, 60°/s) to the start position for the next repetition. Verbal encouragement and real-time visual torque feedback were provided to instruct participants when to contract or relax and to ensure maximal effort. Peak torque personal records were plotted on the graphs to encourage participants to progress the torques produced. Two, three, and four sets per session with rests of 60-90 s were performed in weeks 1, 2, and 3-6, respectively, resulting in an average of 10.5 weekly sets. Sets of 10 CON-RT repetitions were performed first in each session, and the work in each set was then calculated. ECC-RT sets were then performed with sufficient (usually ∼6-8) repetitions to provide the same total work between conditions [20]. Thus, both effort and training volume were matched between conditions although peak torques differed. The torque‒angular displacement integral (i.e., work; J) of the contraction phase of each repetition was calculated using LabChart software (Powerlab System; ADInstruments, Australia) and summed over all sets and sessions to provide the total training volume [25].

### 2.6 Statistical analyses

Normality of data was checked by Shapiro-Wilk’s test, and homogeneity of variances was tested by Levene’s test. Analysis of covariance was used to compare the effects of CON-RT vs. ECC-RT, with the difference between pre- to post-training values inputted as dependent variables, baseline values as covariates (to remove any influence of baseline variance), group as a fixed factor, and region, joint angle, and joint angular velocity as fixed factors for architecture, isometric strength, and dynamic strength test analyses, respectively. Adjusted marginal pre- to post-training mean differences were compared to 0 using paired t-tests to verify the training effect in relation to baseline. Bonferroni correction was applied to multiple-comparison tests. A P value < 0.05 was accepted as statistically significant. Effect sizes (ES) were calculated as the post-training mean minus the pre-training mean, divided by the pooled pre-training standard deviation. ESs of 0.00-0.19 were considered trivial, 0.20-0.49 small, 0.50-0.79 moderate, and ≥ 0.80 large. Pearson’s correlation tests were used to test the linear relationship between outcomes. Correlations (r) of 0.00-0.20 were considered very weak, 0.21-0.40 weak, 0.41-0.60 moderate, 0.61-0.80 strong, and ≥0.81 very strong. The data were expressed as mean, standard deviation, and 95% confidence intervals (CI). Analyses were performed using JASP v.0.16.2 (JASP Team, Netherlands).

## 3 RESULTS

All participants completed all training sessions, and no adverse events were reported during the intervention. Participants’ energy and macronutrient intakes were: 27.1 ± 3.9 kcal/kg/d, protein: 1.6 ± 0.6 g/kg/d, carbohydrate: 3.1 ± 1.2 g/kg/d, and fat: 0.9 ± 0.3 g/kg/d. Total work (training volume) was not different between conditions (CON-RT = 11.9 ± 2.6 vs. ECC-RT = 11.9 ± 2.6 kJ; P = 0.994). No difference was observed for progression in the peak torques achieved across the sessions (slope, P = 0.481), indicating a similar load progression between training modes (+1%/session).

### 3.1 Muscle architecture and size

For FL, a significant condition (P < 0.001) but no region (P = 0.282) or interaction (P = 0.999) effects were detected. No significant changes were detected in CON-RT, but moderate increases were observed in ECC-RT (proximal: CON-RT = -2%; ECC-RT = 5%; distal: CON-RT = 0%; ECC-RT = 7%). For PA, significant condition (P = 0.039) and region (P = 0.024) but no interaction (P = 0.206) effects were detected. Large increases in CON-RT but moderate increases in ECC-RT were observed (proximal: CON-RT = 10%; ECC-RT = 4%; distal: CON-RT = 6%; ECC-RT = 4%). For MT, no significant condition (P = 0.092), region (P = 0.551), or interaction (P = 0.386) effects were observed. Both conditions showed small-to-moderate increases (proximal: CON-RT = 9%; ECC-RT = 9%; distal: CON-RT = 6%; ECC-RT = 11%). Results are illustrated in Figure 2 and presented in Table S1.

**Figure 2.**
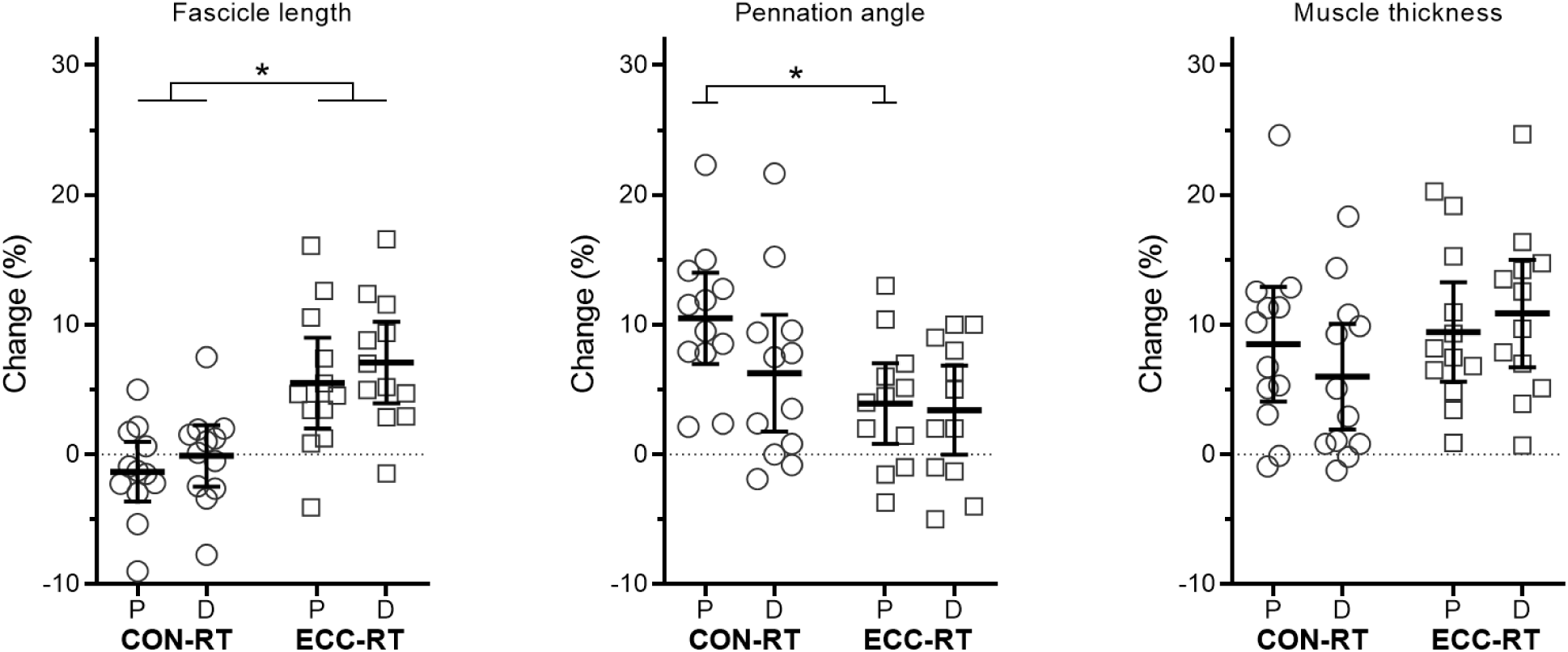
Percentage changes in tibialis anterior muscle architecture at proximal (P) and distal (D) regions after concentric (CON-RT) and eccentric (ECC-RT) resistance training. Data are mean and 95%CI, with open circles and squares representing individual data points. *P < 0.05 difference between conditions.

### 3.2 Maximal voluntary isometric contraction and torque-angle relationship

For maximum isometric torque, no significant condition (P = 0.846), angle (P = 0.363) or interaction (P = 0.278) effects were observed. Both conditions showed trivial-to-moderate increases in the average of the tested angles (CON-RT = 8%, ECC-RT = 8%), with CON-RT significantly improving only at -5°PF and 0°PF while ECC-RT improved at all angles except at 20°PF. The isometric peak torque angle shifted towards shorter muscle lengths for CON-RT (-16%) but did not change for ECC-RT (5%), with a significant difference revealed between conditions (P = 0.046). The torque‒angle plateau region width did not significantly change for CON-RT (-8%) but increased for ECC-RT (12%), with a significant difference revealed between conditions (P = 0.007). Results are illustrated in Figure 3 and presented in Table S2.

**Figure 3.**
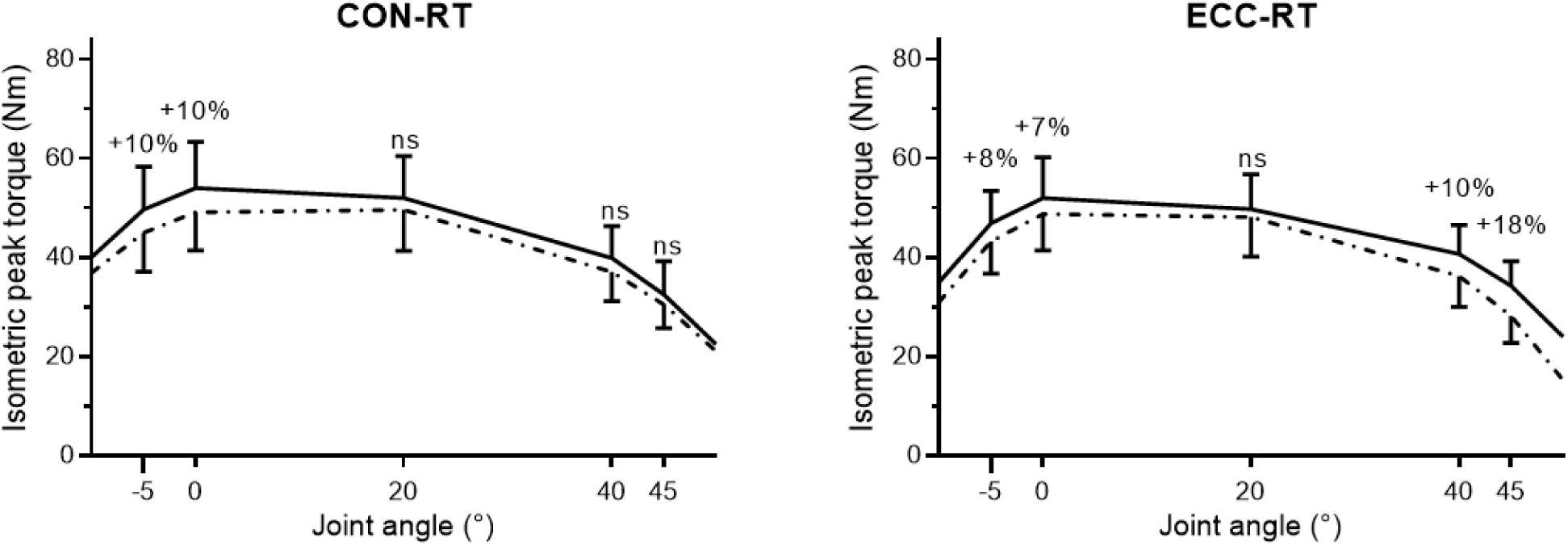
Changes in maximal voluntary isometric contraction torque after concentric (CON-RT) and eccentric (ECC-RT) resistance training according to ankle position (-5°, 0°, 20°, 40°, and 45°-plantarflexion). Data are mean and 95%CI. Dot-dashed line = pre-training. Solid line = post-training. Percentageresults indicate when a significant (P < 0.05) pre- to post-training change occurred. ns = not significant.

### 3.3 Rate of torque development

For RTD in 0-75ms and 0-250ms intervals, no significant condition (P = 0.894, 0.695), angle (P = 0.081, 0.234), or interaction (P = 0.125, 0.080) effects were observed. Both conditions showed moderate increases for the average of the tested angles in both 0-75ms (CON-RT = 24%; ECC-RT = 24%) and 0-250ms (CON-RT = 17%; ECC-RT = 15%) time intervals, but CON-RT significantly improved RTD only at 0°PF, 20°PF, and 40°PF while ECC-RT improved only at 20°PF, 40°PF, and 45°PF. For rRTD in 0-75ms and 0-250ms intervals, no significant condition (P = 0.840, 0.949) or interaction (P = 0.308, 0.178) effects were observed, but a significant effect of angle was observed (P = 0.013, 0.034), with changes at 40°PF and 45°PF being greater than at -5°PF, mainly driven by ECC-RT improvements. Results are illustrated in Figure 4 and presented in Table S3 and Table S4.

**Figure 4.**
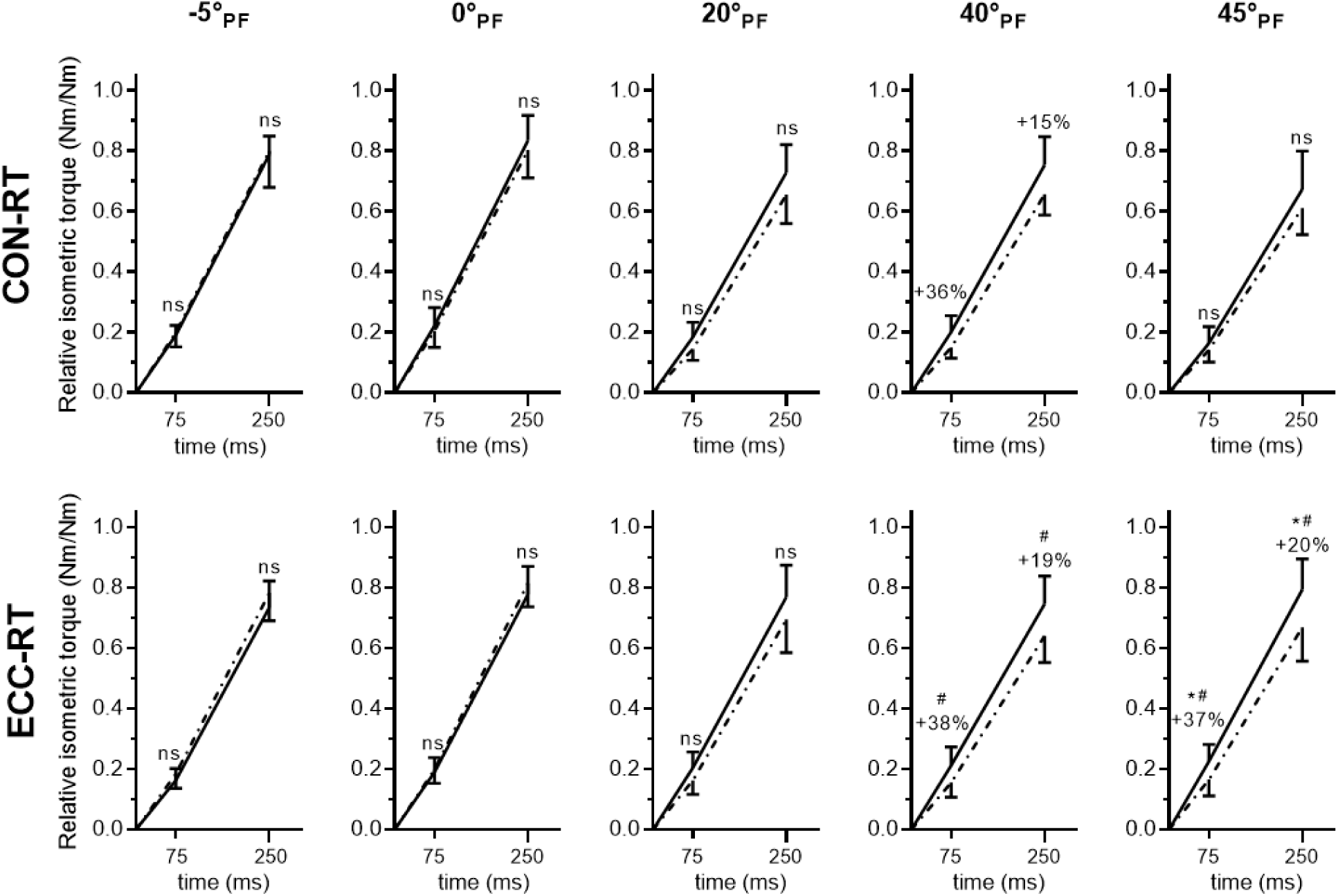
Changes in torque relative to peak torque in the time intervals 0-75ms and 0-250ms, representing the relative rate of torque development after concentric (CON-RT) and eccentric (ECC-RT) resistance training at each ankle position (-5° to 45°-plantarflexion; PF). Data are mean and 95%CI. Dot-dashed line = pre-training. Solid line = post-training. Percentage results indicate when a significant (P < 0.05) pre- to post-training change occurred. ns = not significant. *P < 0.05 difference between conditions for the same angle and time interval. ^#^P < 0.05 greater than -5°PF for the same condition.

### 3.4 Maximal voluntary concentric contraction

For maximum concentric tests either preceded by an isometric or an eccentric contraction, no significant velocity (P = 0.841, 0.506) or interaction (P = 0.970, 0.842) effects were observed, but a significant effect of condition was observed for eccentric-preceded concentric tests (P = 0.047; but not for isometric-preceded tests, P = 0.160), with greater gains observed for ECC-RT. CON-RT showed small increases on isometric-preceded concentric tests (average of the three velocities = 11%) but no significant change after eccentric-preceded concentric tests (8%), whereas ECC-RT showed significant small-to-moderate increases for both isometric-preceded concentric tests (17%) and eccentric-preceded concentric tests (18%). On average, for all angular velocities and contraction histories, changes in concentric strength were significantly greater (P = 0.027) for ECC-RT (17%) than CON-RT (9%). No significant changes in either group and no significant differences between conditions were observed for the estimated maximum shortening velocity in both isometric-preceded (CON-RT = -4%; ECC-RT = 6%; P = 0.495) and eccentric-preceded (CON-RT = -4%; ECC-RT = 12%; P = 0.372) tests. For concentric work done in tests either preceded by an isometric or eccentric contraction, no significant condition (P = 0.922, 0.160), velocity (P = 0.637, 0.612), or interaction (P = 0.603, 0.209) effects were observed; increases in the average concentric work were of small magnitude and similar between conditions (CON-RT = 11%; ECC-RT = 11%). Results for maximum concentric strength at each angular velocity are illustrated in Figure 5 and presented in Table S5.

**Figure 5.**
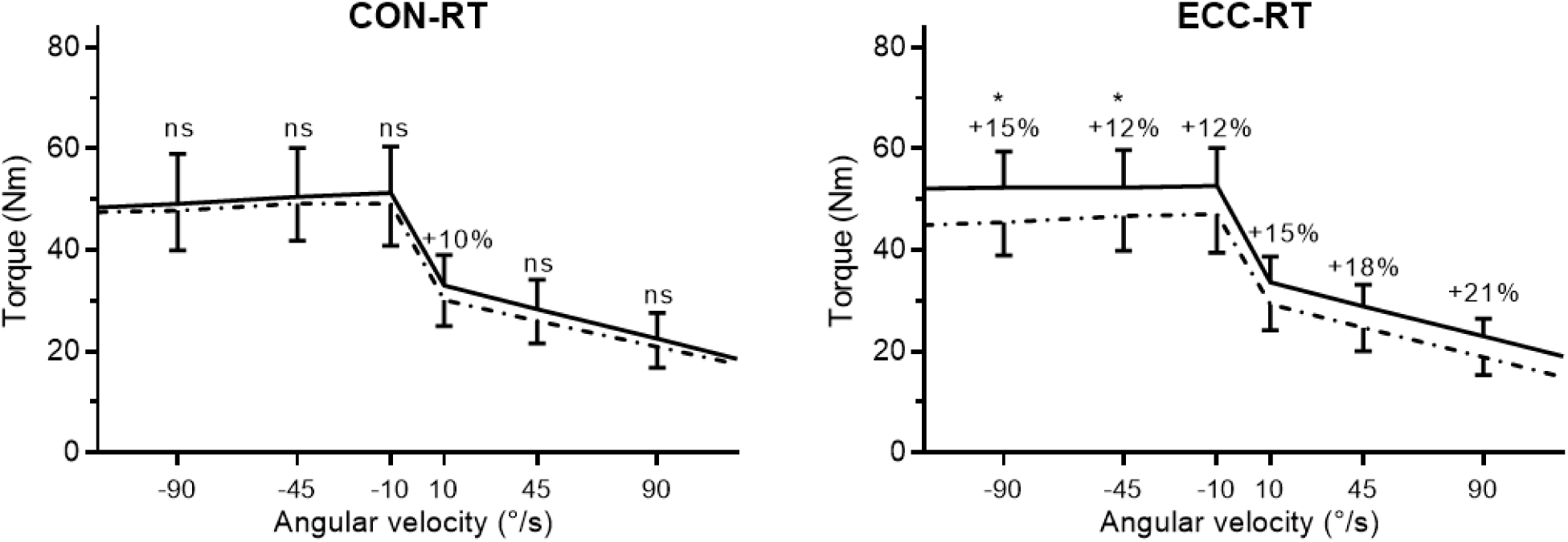
Changes in eccentric and concentric peak torques after concentric (CON-RT) and eccentric (ECC-RT) resistance training at each angular velocity (10°/s, 45°/s, 90°/s; x-axis, negative/positive are eccentric/concentric; results are averaged for both history-of-contraction modes). Dot-dashed line = pre-training. Solid line = post-training. Percentage results indicate when a significant (P < 0.05) pre- to post-training change occurred. ns = not significant. *P < 0.05 difference between conditions at the same velocity.

### 3.5 Maximal voluntary eccentric contraction

For maximum eccentric tests either preceded by an isometric or a concentric contraction, no significant velocity (P = 0.928, 0.804) or interaction (P = 0.539, 0.895) effects were observed, but a significant effect of condition was observed (P < 0.001 both), with greater gains observed for ECC-RT. CON-RT showed no significant changes at any tested angular velocities whereas ECC-RT showed small-to-moderate increases, with significant differences between groups detected at 45°/s and 90°/s. On average, for all angular velocities and contraction histories, changes in eccentric peak torque were significantly greater (P < 0.001) for ECC-RT (13%) than CON-RT (3%). For net eccentric torque (i.e., torque above the isometric maximum), there was a significant moderate reduction in CON-RT but a moderate increase for ECC-RT, with a significant difference between conditions (P < 0.001). In relative terms, net eccentric torque decreased in CON-RT from 16% to 7% (-9%) while it increased in ECC-RT from 15% to 20% (+5%). Results for maximum eccentric strength at each angular velocity are illustrated in Figure 5 and presented in Table S6.

### 3.6 Associations between changes in architecture and changes in function

Given the lack of significant regional training effects on muscle architecture, results are presented without distinguishing between proximal and distal regions, focusing on exploring associations between significant group-level changes in function with changes in architecture after training. When considering group pooled results, significant (P < 0.05), the following correlations were detected: (i) FL was positively correlated with plateau region width (r = 0.54; Figure 6A), and (ii) FL was negatively correlated with rRTD, with magnitude varying across tests and significance detected for contractions at 40°PF 0-75ms (r = -0.41) and 0°PF 0-250ms (r = -0.51; Figure 6B). After CON-RT, the following were detected: (i) FL was positively correlated with isometric peak torque at -5°PF (r = 0.69), and (ii) FL was negatively correlated with relative net eccentric peak torque (r = -0.71). After ECC-RT, the following were detected: (i) PA was positively correlated with dynamic function, with magnitude varying across tests and significance detected for average concentric peak torque in isometric- and eccentric-preceded tests at 10°/s (r = 0.64; Figure 6C), concentric work in the three isometric-preceded tests (10°/s, r = 0.58; 45°/s, r = 0.58; 90°/s, r = 0.58; and the average of them, r = 0.63), eccentric peak torque in isometric- and concentric-preceded tests at 10°/s (r = 0.61; r = 0.58, respectively; and the average of them, r = 0.61), and average eccentric peak torque in isometric- and concentric-preceded tests in all velocities (r = 0.58; Figure 6D); (ii) FL was negatively correlated with rRTD at 40°PF 0-75ms (r = -0.66) and 40°PF 0-250ms (r = - 0.63); and (iii) PA was positively correlated with rRTD at 0°PF 0-75ms (r = 0.58) and 40°PF 0-75ms (r = 0.67). Scatter plots with the main significant correlations are presented in Figure 6.

**Figure 6.**
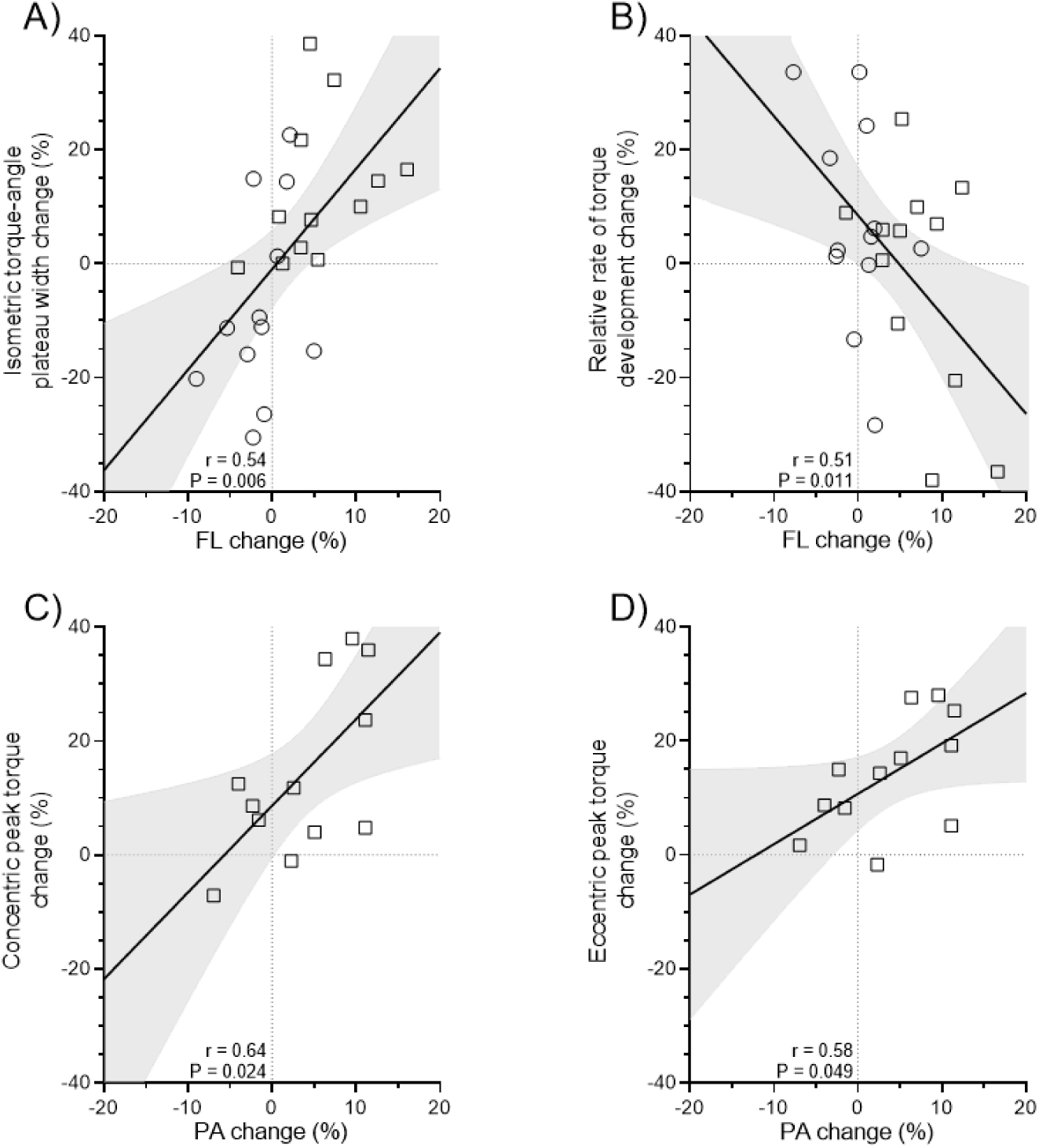
Associations between changes in muscle architecture and changes in muscle function. A) Changes in fascicle length (FL) and changes in isometric torque-angle plateau region width. B) Changes in FL and relative rate of torque development at 0°PF 0-250ms. C) Changes in pennation angle (PA) and changes in average concentric peak torque in the isometric- and eccentric-preceded tests at 10°/s. D) Changes in PA and changes in average eccentric peak torque in the isometric- and concentric-preceded tests at 10°/s, 45°/s, and 90°/s. Data are represented by the regression line with 95%CI, with open circles and squares representing individual data points from CON-RT and ECC-RT conditions, respectively.

## 4 DISCUSSION

This study offers a novel perspective by comparing the effects of CON-RT and ECC-RT on regional muscle architecture and functional parameters under an effort- and volume-equated design. Both training modes increased MT similarly, but architectural adaptations were training mode-specific, with CON-RT promoting greater PA increase and ECC-RT promoting greater FL increase. Importantly, changes in function were less associated with the contraction mode (or velocity or angle) used in training and more associated with specific architectural adaptations. Thus, adaptation specificity predominated over training specificity.

### 4.1 Muscle architecture and size

Previous studies have shown mixed results regarding the relative hypertrophic effects of CON-RT versus ECC-RT. However, the literature consistently indicates that when training volume is equated, both modalities produce similar increases in muscle size [20,21,25,26], a finding confirmed by the present study. However, unlike previous research on other lower-limb muscles, such as vastus lateralis [7,10,16], no condition-dependent region-specific growth was found. This is because regional hypertrophy may be more pronounced in muscles with greater architectural variation along their length (e.g., in vastus lateralis [12]), where different regions experience distinct mechanical stimuli regardless of the whole-muscle stimulus imposed [27].

Importantly, MT changes can result from alterations in either PA or FL due to their trigonometric relationship [28]. The present study provides the first evidence that muscle architecture adaptations differ between effort- and volume-equated ECC-RT and CON-RT. Such contraction mode-specific adaptive responses, with PA increasing more in CON-RT (8%) than ECC-RT (4%) and FL increasing more in ECC-RT (6%) than CON-RT (-5%), are consistent with many [8,10,11,16], but not all [7,29], previous studies. Additionally, the average relative increase in MT was significantly correlated with changes in both PA and FL (r = 0.54, 0.51, respectively; data not shown). These findings indicate that, despite producing contraction mode-specific architectural adaptations, overall muscle hypertrophy remained similar [8,11,16], suggesting that both CON-RT and ECC-RT effectively enhance muscle size through distinct changes in muscle architecture.

Exercise-induced PA increases are considered to primarily result from parallel myofibril addition [2,6], where myofibrillogenesis increases radial fiber size causing bulging and lateral displacement of fiber bundles. However, increases in sarcoplasmic protein and intramuscular fluid content may also enlarge fascicles [28,30]. That is, increased PA represents more than just parallel sarcomere addition. Although fluid and connective tissues significantly impact muscle function [31,32], the extent to which training-induced sarcoplasm enlargement contributes to muscle function and growth remains underexplored [28], particularly in response to different training modalities such as CON-RT and ECC-RT. CON-RT may speculatively induce disproportionally greater sarcoplasmic hypertrophy due to its higher metabolic cost [33], as suggested to occur in very-high-volume traditional training [30], potentially enhancing intracellular glycogen storage and thus intracellular water, since 3-4 grams of water bind to each gram of glycogen in skeletal muscle. This could help explain the greater increases in PA often observed after CON-RT for a given level of myofibrillar hypertrophy. However, this remains speculative and warrants further investigation.

Regarding increased FL, this is often attributed to serial sarcomere addition within the fibers [6,34,35], although increases in resting sarcomere length [36] and longitudinal sliding of parallel intrafascicular-terminating fibers [32] may also contribute. That is, increased FL represents more than just serial sarcomere addition. To date, only two training studies [36,37] in humans have directly assessed changes in both sarcomere and fascicle lengths to estimate sarcomere number *in vivo*, rather than drawing assumptions from FL changes alone. Three weeks of Nordic hamstring exercise ECC-RT increased sarcomere length [36] but nine weeks of training apparently increased sarcomere number [37] of the biceps femoris long head. Since substantial FL changes were reported in both studies [36,37], these data suggest that FL changes may not always prove a useful proxy for serial sarcomere number changes [38], and may not always allow precise inferences about muscle function. Thus, detailed functional assessments are necessary.

### 4.2 Voluntary isometric contraction and the torque‒angle relationship

For isometric strength, if training specificity predominated, ECC-RT might produce greater gains due to the higher training torques, with both training modes improving strength primarily in the middle of the range of motion where peak torques occurred. By contrast, if structural adaptations predominated, regardless of training mode, one might expect: i) increased PA would enhance muscle force capacity through increased physiological cross-sectional area, and ii) assuming that exercise-induced FL and torque-angle changes result from changes in both serial sarcomere number and the length‒tension relationship [1,3], an increase in FL should shift the peak torque angle toward longer lengths (greater dorsiflexion) and broaden the torque‒angle relationship whereas FL decrease would result in opposite outcomes [1,3].

In the present study, overall increases in isometric peak torque were similar between conditions. Since cross-sectional area was not measured and thus physiological cross-sectional area, or functional cross-sectional area, could not be calculated to better understand the mechanical advantage of PA in force production [2], the calculation using muscle thickness presents an alternative; MT×sec(PA) [39]. Post-hoc calculations (data not shown) indicated that CON-RT and ECC-RT had similar increases in functional muscle thickness (CON-RT = 7%; ECC-RT = 9%; P = 0.241) and relative pennation mechanical advantage (calculated as FMT/MT [2]; CON-RT = 0.4%; ECC-RT = 0.2%; P = 0.084), partially explaining the similar overall isometric peak torque results (CON-RT = 8%; ECC-RT = 8%).

Despite the similar overall effects, each training modality produced distinct changes in the torque‒ angle relationship. After CON-RT, the lack of significant change in FL was observed alongside significant increases in isometric peak torque only at shorter muscle lengths, resulting in a leftward shift in the torque‒ angle relation (-2.2°) and slight torque‒angle curve narrowing (-8%, P = 0.059). In contrast, ECC-RT increased FL but isometric peak torques increased at both shorter and longer muscle lengths (but not in the mid-range of the contraction), resulting in a significant plateau region broadening (12%) without peak torque‒angle change. Across both conditions, FL changes were moderately associated with plateau width changes (r = 0.54; Figure 6A) but not with shifts in peak torque angle. Interestingly, in CON-RT, FL increases were positively correlated with higher isometric peak torque at shorter muscle lengths. However, this correlation was driven by a single participant who showed the largest FL increase (7%) and peak torque improvement at shorter lengths (37%) after CON-RT (scatterplot presented in the supplementary material). This participant initially had the lowest peak torque at -5°PF (-21% of optimal angle torque) and showed the largest decrease in torque at longer lengths after training (-16%) – a pattern consistent with increased sarcomere length and sarcomere number reduction. For this participant, fibers may have initially operated at very short lengths at -5°PF, with FL increases allowing fibers to function closer to their optimal length, while potentially reaching passive insufficiency at 45°PF after training. These results highlight that FL increases do not consistently produce torque‒angle rightward shifts, suggesting that different mechanisms may have driven FL adaptations and that functional testing may help infer the microstructural adaptations. Moreover, although FL increases often predominate after ECC-RT, such adaptations are not exclusive to eccentric training, as shown in other studies [7,8,10,16]. Notably, if fascicle adaptations resulted only from serial sarcomere number regulation, FL should have decreased after CON-RT (at least according to present theories [1,3]) and ECC-RT should have clearly shifted the peak torque angle towards longer muscle lengths. However, neither outcome was observed. Thus, the results provide moderate support for the theory that changes in isometric force characteristics aligned more clearly with structural adaptation specificity than contraction mode (or range of motion) specificity, although it is not completely clear which sarcomeric-level changes underpin the structural changes.

### 4.3 Voluntary dynamic contraction

Training-specificity theory predicts that peak torques in each contraction mode improve most after each specific training mode and at the trained velocity. Conversely, structural specificity theory predicts that FL increases, assuming they reflect serial sarcomere addition, should enhance both eccentric and concentric function regardless of training mode, as increasing sarcomere number reduces the per-sarcomere strain and velocity, optimizing force production at non-zero velocities [1,3]. Additionally, increased PA may reflect enhanced parallel hypertrophy and lead to higher architectural gear ratios (Δmuscle length / Δfascicle length) during contraction, allowing more optimal fiber lengths to be adopted and slower fiber velocities to be realized during force production [31,40].

#### 4.3.1 Concentric function and torque‒velocity relationship

In the present study, concentric peak torque increased more in ECC-RT (17%) than CON-RT (9%). Although this result contradicts the specificity principle, CON-RT and ECC-RT often produce similar concentric strength improvements [23], which in itself contradicts the specificity principle; nonetheless, an advantage of ECC-RT has been reported in other studies [23]. While one could suggest interlimb cross-over effects might have influenced the outcomes, this is unlikely as ECC-RT would have been expected to provide a greater cross-over effect than CON-RT [41], but no evidence of this was found. When both limbs are trained, competing neural demands are hypothesized to inhibit inter-limb signaling and thus the typical cross-over effect [42]. Supporting structural specificity, changes in concentric function after ECC-RT were significantly correlated with increased PA (Figure 6C). The greater concentric peak torque with ECC-RT may be explained by improved torque at long muscle lengths during isometric-preceded concentric tests, increased fascicle rotation and thus gear ratio maintaining initial concentric torque [40], and potentially energy storage-release from series elastic tissues during eccentric-preceded concentric tests, with stored energy in the eccentric phase assisting concentric force production [43]. This is consistent with concentric torque improvements being greatest for ECC-RT at the higher speed tests (45°s and 90°/s), where stretch-shorten cycle efficiency should be amplified [43], and least in 10°/s and isometric-preceded tests (based on the ESs; see Table S4). However, concentric work improvements averaged across the six concentric tests (three velocities with two contraction histories) were similar between conditions. The observed differences in concentric torque, but not work, may reflect greater force production at shorter muscle lengths after CON-RT, offsetting the greater force at longer lengths after ECC-RT, resulting in comparable total work.

Additionally, longitudinal muscle growth theoretically increases maximum shortening velocity in zero-load contractions [1,3]. Although ECC-RT showed relatively greater concentric peak torque improvement at 90°/s (21%) than at 10°/s (15%), suggesting a small torque‒velocity relationship shift counter to training specificity, no significant changes in estimated maximum shortening velocity were detected in any condition, and the effect sizes of the changes were small regardless of whether estimates were derived from isometric-preceded or eccentric-preceded concentric strength tests. Although previous studies have reported longer fascicles in faster athletes (e.g., sprint runners [44]) evidence linking changes in FL to changes in high-speed force production or maximum unloaded muscle shortening speed remains scarce. A recent review, for example, reported no direct evidence from longitudinal (training) studies that longitudinal fiber growth increases maximum velocity in either animal or human data [3]. Thus, it has yet to be determined whether training-induced changes in FL directly affect high-speed muscle force capacity.

#### 4.3.2 Eccentric function

In the present study, eccentric peak torque increased more in ECC-RT (13%) than in CON-RT (3%), consistent with the training specificity principle. However, ECC-RT produced similar relative changes in eccentric and concentric peak torques, in contrast with specificity principles. Increases in FL and PA with training may enhance eccentric force production through reduced per-sarcomere lengthening during contraction. However, more recent perspectives suggest cross-bridge theory alone cannot fully explain eccentric strength capacity, with other structural adaptations accounting for the increases in eccentric strength, such as increased titin stiffness (and other series elastic components) enhancing resistance to active lengthening [24,33,45]. Unlike concentric contractions, muscle activation remains unchanged with increasing torque during lengthening, suggesting a significant role of non-contractile tissues in eccentric force production [24,33,45].

Interestingly, in the present study, the concept of ‘net eccentric torque’ (change in peak torque from the isometric to eccentric phase in isometric-preceded tests) was introduced to determine whether eccentric strength changes stemmed from changes in preceding isometric torque or active lengthening itself. After training, net eccentric torque increased only after ECC-RT (5%) while decreasing after CON-RT (-9%), highlighting the capacity of ECC-RT to enhance torque during active lengthening [24,33,45]. Moreover, after CON-RT, strong inverse correlations were observed between changes in FL and net eccentric torque (r = -0.71). The participant mentioned earlier (see section 4.2) with the largest FL increase after CON-RT also showed the most pronounced reductions in eccentric torque (-16%) and net eccentric torque (-48%), suggesting that increases in FL without the specific adaptations to series elastic components (typically seen only after ECC-RT [46]) may impair eccentric strength. Speculatively, if sarcomeres lengthen at rest (as observed previously [36]) without changes in sarcomere number or titin stiffness (to maintain optimal passive force contribution [47]), expected passive tension increases may not occur. Since series elastic structures significantly contribute to eccentric force production [24,33,45], this could explain the inverse relationship between FL increases and eccentric torque reductions after CON-RT. The effects of increased muscle fiber length (either via increased sarcomere number or length) in the presence or absence of increased stiffness of the series elastic components (mainly titin) on muscle function warrant further investigation.

Increased PA may be associated with enhanced maximum strength either through the parallel sarcomere addition that largely underlies increased PA [6] or improved muscle gearing that can result from it [31]. In the present study, PA changes were correlated with improved dynamic function after ECC-RT (Figure 6C-D) but not CON-RT, which showed greater changes in PA. This suggests CON-RT may have induced sarcoplasmic hypertrophy, which does not directly enhance force production. Additionally, muscle gearing is influenced by intramuscular connective tissue properties, particularly the extracellular matrix, which may have undergone remodeling only after ECC-RT [46]. Eng et al. [31] proposed that muscle gearing depends on the ability of connective tissues to support fascicle rotation during high-force contractions, effectively reducing fascicle strain and allowing them to remain closer to optimum length and to operate at a slower shortening speed through the range of motion. Furthermore, extracellular matrix adaptations may improve the sliding capacity of intramuscular collagenous tissues, enabling more efficient fascicle rotation [32]. These mechanisms remain speculative but could explain why PA increases were functionally beneficial after ECC-RT but not CON-RT, as only ECC-RT may have induced the necessary connective tissue adaptations.

### 4.4 Rate of torque development

According to the structural specificity hypothesis, increased PA could enhance RTD through improved muscle gearing, allowing greater force for a given muscle belly shortening during fixed-end contractions [4], while increased FL might reduce RTD by increasing series compliance [18,48]. In the present study, both conditions improved RTD at 0-75ms and 0-250ms time intervals. CON-RT showed significant increases at 0°PF, 20°PF, and 40°PF while ECC-RT showed significant increases at 20°PF, 40°PF, and 45°PF. However, when adjusting for peak torque (relative RTD [rRTD]), only effects at longer muscle lengths in each group remained significant, suggesting that the RTD increases at shorter lengths primarily reflected enhanced force capacity rather than improved contractile velocity [18,48].

While some correlations emerged between PA and rRTD changes, results were inconsistent across angles, time intervals, and groups, preventing clear conclusions. This may be because the tibialis anterior has a small resting PA (thus, gearing effects are less pronounced) [4,31,40], and other adaptations that influence RTD may have also occurred with the training [18]. Future studies should investigate the role of training-induced changes in architectural gear ratios on muscle function, especially in muscles with greater PA. In contrast, negative correlations existed between FL and rRTD changes for both groups, particularly for ECC-RT, which showed greater rRTD improvements than CON-RT. While most participants improved rRTD, those with the largest FL increases showed reduced rRTD effects (Figure 6B), hinting at an effect of increased in-series compliance – a phenomenon that has been observed also in the knee extensors [48]. At the group level, however, FL increases were insufficient to reduce rRTD. It is possible that passive tension, which increases at longer muscle lengths and contributes to enhanced sensory inhibitory feedback [49], was reduced after training. This may have attenuated inhibitory afferent feedback at longer lengths, allowing for rRTD improvements; such a hypothesis remains to be explicitly studied in future research. Regardless, a greater decrease in passive muscle stiffness at longer lengths after ECC-RT might explain the greater rRTD improvement. These and other factors may have counterbalanced the negative effects of increasing FL.

### 4.5 Conclusions

CON-RT and ECC-RT produced similar increases in MT but different architectural and functional adaptations. CON-RT resulted in greater increases in PA and improved peak torque at shorter muscle lengths, resulting in a leftward shift in the torque‒angle relation and slight torque‒angle curve narrowing, but reduced net eccentric peak torque. In contrast, ECC-RT resulted in greater increases in FL, improved peak torque at both shorter and longer muscle lengths (broadening the isometric torque‒angle plateau region), greater increases in rRTD at longer muscle lengths, and greater concentric and eccentric strength gains. These results are contrary to the training-specificity principles. Moreover, significant correlations were identified between changes in muscle architecture (FL and PA) and changes in isometric, concentric, and eccentric strength, and rRTD. These findings indicate that, under the current conditions, functional adaptations were more closely related to specific architectural changes evoked by CON-RT and ECC-RT than to training specificities. Muscle function seems to adapt to the structural changes induced by training, regardless of the training program used. Thus, training programs may be designed with a focus on evoking specific structural adaptations, which may provide functional benefits that do not necessarily follow principles of training specificity.

## 5 PERSPECTIVES

Both training modes can be implemented to increase muscle size, but ECC-RT may be preferred for improving muscle function across a wider range of tasks. While the results are specific to ankle-dorsiflexion training, similar architectural responses in other muscles (e.g., knee extensors and flexors [8,10,11,16]) support broader applicability, assuming functional changes follow architectural adaptations [1,5]. However, extrapolation requires caution since architecture‒function associations are not always strong, and macro-level adaptations in PA and FL may not fully capture the underlying microstructural changes that ultimately influence function. Additionally, although neural adaptations should also be considered, a recent study demonstrated that including architectural variables in models predicting isometric and dynamic strength gains after isoinertial knee-extension training improved those models’ predictive power [50]. This highlights the effect of architecture on function and the need for further investigations into the multiple factors driving changes in muscle function. Future studies should incorporate dynamic architecture analyses during contractions to explore the role of architecture in function beyond what is captured with resting architecture measurements [31], and explore micro- and macrostructural muscular adaptations to better understand the association between these responses and changes in muscle function (regardless of whether outcomes at different structural levels align or not).

## Data Availability

All data produced in the present study are available upon reasonable request to the authors

## Acknowledgments

We would like to thank Edith Cowan University for the PhD HDR scholarship provided to JPN.

## Author contributions

JPN collected and analyzed the data and wrote the first draft. KN and AJB provided supervision and made substantial contributions to the original work.

