## supplementary material for "The training specificity versus structural adaptation paradox: Differential effects of isokinetic concentric and eccentric resistance training on muscle architecture and function in young men"

**Table S1.** Training effects on resting architecture parameters.

| | Pre | Post | ES |
|---|---|---|---|
| Fascicle length - proximal (mm) | | | |
| CON-RT | 58.2 ± 7.7 | 57.3 ± 8.2 | -0.12 |
| ECC-RT | 57.5 ± 7.6 | 60.6 ± 8.4*# | 0.42 |
| Fascicle length - distal (mm) | | | |
| CON-RT | 58.8 ± 9.3 | 58.8 ± 9.8 | -0.01 |
| ECC-RT | 57.0 ± 8.1 | 61.0 ± 8.4*# | 0.46 |
| Pennation angle - proximal (°) | | | |
| CON-RT | 12.5 ± 1.0 | 13.8 ± 1.0*# | 1.37 |
| ECC-RT | 12.9 ± 0.9 | 13.4 ± 0.8* | 0.50 |
| Pennation angle - distal (°) | | | |
| CON-RT | 11.7 ± 0.6 | 12.4 ± 0.9* | 1.07 |
| ECC-RT | 12.1 ± 0.7 | 12.5 ± 0.8* | 0.64 |
| Muscle thickness - proximal (mm) | | | |
| CON-RT | 12.6 ± 2.0 | 13.6 ± 1.8* | 0.57 |
| ECC-RT | 12.8 ± 1.6 | 14.0 ± .17* | 0.68 |
| Muscle thickness - distal (mm) | | | |
| CON-RT | 12.0 ± 1.9 | 12.6 ± 2.0* | 0.40 |
| ECC-RT | 11.9 ± 1.6 | 13.2 ± 1.9* | 0.74 |

**Notes:** CON-RT = concentric resistance training; ECC-RT = eccentric resistance training; ES = effect size; *P < 0.05 vs. Pre. #P < 0.05 greater effect than the other condition.

**Table S2.** Training effects on dorsiflexion isometric strength parameters.

| | Pre | Post | ES |
|---|---|---|---|
| Isometric PT -5°$_{PF}$ (Nm) | | | |
| CON-RT | 45.0 ± 12.4 | 49.6 ± 13.7* | 0.41 |
| ECC-RT | 43.2 ± 10.3 | 46.5 ± 10.7* | 0.29 |
| Isometric PT 0°$_{PF}$ (Nm) | | | |
| CON-RT | 49.1 ± 12.2 | 54.0 ± 14.6* | 0.42 |
| ECC-RT | 48.7 ± 11.5 | 52.3 ± 12.9* | 0.31 |
| Isometric PT 20°$_{PF}$ (Nm) | | | |
| CON-RT | 49.6 ± 12.9 | 52.4 ± 13.3 | 0.23 |
| ECC-RT | 48.1 ± 12.5 | 49.7 ± 11.1 | 0.13 |
| Isometric PT 40°$_{PF}$ (Nm) | | | |
| CON-RT | 37.1 ± 9.3 | 39.9 ± 10.1 | 0.30 |
| ECC-RT | 36.2 ± 9.8 | 39.7 ± 9.1* | 0.38 |
| Isometric PT 45°$_{PF}$ (Nm) | | | |
| CON-RT | 30.5 ± 7.6 | 33.0 ± 10.6 | 0.31 |
| ECC-RT | 28.2 ± 8.5 | 33.3 ± 7.8* | 0.64 |
| Average isometric PT (Nm) | | | |
| CON-RT | 42.3 ± 10.6 | 45.8 ± 12.2* | 0.35 |
| ECC-RT | 40.9 ± 10.0 | 44.3 ± 9.9* | 0.34 |
| Angle of isometric PT (°) | | | |
| CON-RT | 12.2 ± 4.8 | 10.1 ± 4.2*# | -0.51 |
| ECC-RT | 11.8 ± 3.8 | 12.4 ± 4.1 | 0.14 |
| Width of plateau region (°) | | | |
| CON-RT | 16.1 ± 2.9 | 14.9 ± 3.0 | -0.48 |
| ECC-RT | 15.0 ± 2.5 | 16.8 ± 1.9*# | 0.66 |

**Notes:** CON-RT = concentric resistance training; ECC-RT = eccentric resistance training; PT = peak torque; PF = plantarflexion; ES = effect size; *$P < 0.05$ vs. Pre. #$P < 0.05$ greater effect than the other condition.

**Table S3.** Training effects on dorsiflexion rate of torque development parameters.

| | Pre | Post | ES |
|---|---|---|---|
| ***0-75ms*** | | | |
| RTD -5°$_{PF}$ (Nm/s) | | | |
| CON-RT | 95.9 ± 41.5 | 106.4 ± 39.6 | 0.28 |
| ECC-RT | 82.5 ± 33.1 | 79.1 ± 34.1 | -0.09 |
| RTD 0°$_{PF}$ (Nm/s) | | | |
| CON-RT | 113.9 ± 49.4 | 134.2 ± 47.5* | 0.39 |
| ECC-RT | 114.0 ± 55.2 | 117.2 ± 58.9 | 0.06 |
| RTD 20°$_{PF}$ (Nm/s) | | | |
| CON-RT | 84.6 ± 36.2 | 113.7 ± 45.6* | 0.64 |
| ECC-RT | 94.8 ± 54.2 | 123.6 ± 51.8* | 0.63 |
| RTD 40°$_{PF}$ (Nm/s) | | | |
| CON-RT | 67.5 ± 22.0 | 96.8 ± 31.0* | 1.31 |
| ECC-RT | 67.8 ± 23.9 | 104.3 ± 32.3*$^{§}$ | 1.62 |
| RTD 45°$_{PF}$ (Nm/s) | | | |
| CON-RT | 53.9 ± 23.0 | 63.3 ± 22.9 | 0.34 |
| ECC-RT | 58.9 ± 32.9 | 95.9 ± 41.5*$^{\#§}$ | 1.33 |
| ***0-250ms*** | | | |
| RTD -5°$_{PF}$ (Nm/s) | | | |
| CON-RT | 122.5 ± 36.4 | 138.1 ± 41.4 | 0.49 |
| ECC-RT | 115.5 ± 26.8 | 116.5 ± 30.8 | 0.03 |
| RTD 0°$_{PF}$ (Nm/s) | | | |
| CON-RT | 143.1 ± 34.5 | 164.2 ± 33.2* | 0.57 |
| ECC-RT | 144.4 ± 40.5 | 148.5 ± 47.6 | 0.11 |
| RTD 20°$_{PF}$ (Nm/s) | | | |
| CON-RT | 120.8 ± 36.3 | 143.3 ± 37.8* | 0.54 |
| ECC-RT | 126.0 ± 47.9 | 144.4 ± 40.2* | 0.44 |
| RTD 40°$_{PF}$ (Nm/s) | | | |
| CON-RT | 93.1 ± 21.6 | 114.4 ± 22.0* | 1.06 |
| ECC-RT | 87.7 ± 19.1 | 116.5 ± 29.8*$^{§}$ | 1.43 |
| RTD 45°$_{PF}$ (Nm/s) | | | |
| CON-RT | 71.9 ± 17.2 | 82.5 ± 22.0 | 0.42 |
| ECC-RT | 74.3 ± 32.5 | 104.9 ± 32.2*$^{\#§}$ | 1.20 |

**Notes:** CON-RT = concentric resistance training; ECC-RT = eccentric resistance training; RTD = rate of torque development; PT = peak torque; PF = plantarflexion; ES = effect size; *P < 0.05 vs. Pre. $^{\#}$P < 0.05 greater than the other condition. $^{§}$P < 0.05 greater than -5°$_{PF}$ for the same condition.

**Table S4.** Training effects on dorsiflexion relative rate of torque development parameters.

| | Pre | Post | ES |
|---|---|---|---|
| ***0-75ms*** | | | |
| rRTD -5°$_{PF}$ (Nm/s/Nm) | | | |
| CON-RT | 2.3 ± 1.0 | 2.2 ± 0.7 | -0.08 |
| ECC-RT | 2.0 ± 1.0 | 1.8 ± 0.8 | -0.26 |
| rRTD 0°$_{PF}$ (Nm/s/Nm) | | | |
| CON-RT | 2.4 ± 1.1 | 2.7 ± 1.2 | 0.26 |
| ECC-RT | 2.4 ± 1.0 | 2.3 ± 1.0 | -0.11 |
| rRTD 20°$_{PF}$ (Nm/s/Nm) | | | |
| CON-RT | 1.8 ± 0.8 | 2.3 ± 1.0 | 0.61 |
| ECC-RT | 2.0 ± 1.0 | 2.6 ± 1.1 | 0.66 |
| rRTD 40°$_{PF}$ (Nm/s/Nm) | | | |
| CON-RT | 1.9 ± 0.8 | 2.6 ± 1.1* | 0.80 |
| ECC-RT | 2.0 ± 1.0 | 2.8 ± 1.2*$^{§}$ | 0.87 |
| rRTD 45°$_{PF}$ (Nm/s/Nm) | | | |
| CON-RT | 1.9 ± 0.8 | 2.2 ± 1.1 | 0.31 |
| ECC-RT | 2.2 ± 1.1 | 3.0 ± 1.2*$^{\#§}$ | 0.82 |
| ***0-250ms*** | | | |
| rRTD -5°$_{PF}$ (Nm/s/Nm) | | | |
| CON-RT | 2.8 ± 0.7 | 2.8 ± 0.4 | -0.03 |
| ECC-RT | 2.7 ± 0.6 | 2.6 ± 0.6 | -0.28 |
| rRTD 0°$_{PF}$ (Nm/s/Nm) | | | |
| CON-RT | 3.0 ± 0.7 | 3.1 ± 0.6 | 0.27 |
| ECC-RT | 3.0 ± 0.5 | 2.8 ± 0.6 | -0.24 |
| rRTD 20°$_{PF}$ (Nm/s/Nm) | | | |
| CON-RT | 2.5 ± 0.6 | 2.8 ± 0.6 | 0.48 |
| ECC-RT | 2.6 ± 0.7 | 2.9 ± 0.7 | 0.50 |
| rRTD 40°$_{PF}$ (Nm/s/Nm) | | | |
| CON-RT | 2.6 ± 0.5 | 3.0 ± 0.6* | 0.77 |
| ECC-RT | 2.5 ± 0.6 | 3.0 ± 0.7*$^{§}$ | 0.91 |
| rRTD 45°$_{PF}$ (Nm/s/Nm) | | | |
| CON-RT | 2.4 ± 0.6 | 2.7 ± 0.8 | 0.39 |
| ECC-RT | 2.6 ± 0.7 | 3.2 ± 0.7*$^{\#§}$ | 0.86 |

**Notes:** CON-RT = concentric resistance training; ECC-RT = eccentric resistance training; rRTD = relative rate of torque development; PF = plantarflexion; ES = effect size; *$P < 0.05$ vs. Pre. $^{\#}P < 0.05$ greater than the other condition. $^{§}P < 0.05$ greater than -5°$_{PF}$ for the same condition.

**Table S5.** Training effects on dorsiflexion concentric strength parameters.

| | Pre | Post | ES |
|---|---|---|---|
| ***Preceded by an isometric contraction*** | | | |
| Concentric PT 10°/s (Nm) | | | |
| CON-RT | 29.0 ± 6.9 | 32.4 ± 10.1* | 0.46 |
| ECC-RT | 28.5 ± 7.9 | 32.9 ± 7.9* | 0.61 |
| Concentric PT 45°/s (Nm) | | | |
| CON-RT | 24.4 ± 6.2 | 26.8 ± 9.4 | 0.38 |
| ECC-RT | 23.6 ± 6.6 | 27.0 ± 6.2* | 0.55 |
| Concentric PT 90°/s (Nm) | | | |
| CON-RT | 18.8 ± 6.0 | 20.9 ± 7.6 | 0.36 |
| ECC-RT | 17.5 ± 5.5 | 21.0 ± 5.1* | 0.62 |
| Average concentric PT (Nm) | | | |
| CON-RT | 24.1 ± 6.3 | 26.7 ± 8.9* | 0.42 |
| ECC-RT | 23.2 ± 6.5 | 27.0 ± 6.2* | 0.61 |
| Estimated concentric Vmax (°/s) | | | |
| CON-RT | 244.6 ± 74.4 | 232.8 ± 44.5 | -0.19 |
| ECC-RT | 224.4 ± 51.9 | 241.4 ± 61.0 | 0.27 |
| Concentric work 10°/s (J) | | | |
| CON-RT | 21.0 ± 5.6 | 24.2 ± 8.5* | 0.56 |
| ECC-RT | 21.2 ± 5.7 | 24.3 ± 6.4* | 0.55 |
| Concentric work 45°/s (J) | | | |
| CON-RT | 19.2 ± 4.5 | 21.7 ± 8.1* | 0.51 |
| ECC-RT | 19.1 ± 5.1 | 20.8 ± 5.7 | 0.36 |
| Concentric work 90°/s (J) | | | |
| CON-RT | 19.0 ± 4.2 | 20.3 ± 6.4 | 0.32 |
| ECC-RT | 18.3 ± 4.2 | 20.4 ± 4.5* | 0.52 |
| ***Preceded by an eccentric contraction*** | | | |
| Concentric PT 10°/s (Nm) | | | |
| CON-RT | 31.1 ± 9.2 | 33.5 ± 9.1 | 0.28 |
| ECC-RT | 30.2 ± 8.4 | 34.4 ± 8.4* | 0.48 |
| Concentric PT 45°/s (Nm) | | | |
| CON-RT | 27.7 ± 8 | 29.7 ± 9.1 | 0.26 |
| ECC-RT | 25.9 ± 8.2 | 31.1 ± 7.2* | 0.66 |
| Concentric PT 90°/s (Nm) | | | |
| CON-RT | 23.1 ± 7.5 | 24.8 ± 8.4 | 0.27 |
| ECC-RT | 20.3 ± 6.6 | 24.6 ± 6.3* | 0.66 |
| Average concentric PT (Nm) | | | |
| CON-RT | 27.3 ± 8.1 | 29.4 ± 8.8 | 0.31 |
| ECC-RT | 25.5 ± 7.2 | 30.0 ± 6.8* | 0.62 |
| Estimated concentric Vmax (°/s) | | | |
| CON-RT | 348.5 ± 96.1 | 335.4 ± 102.4 | -0.12 |
| ECC-RT | 309.4 ± 119.8 | 345.4 ± 112.0 | 0.33 |
| Concentric work 10°/s (J) | | | |
| CON-RT | 24.0 ± 7.1 | 26.7 ± 7.9* | 0.44 |
| ECC-RT | 24.2 ± 5.3 | 25.9 ± 5.9* | 0.28 |
| Concentric work 45°/s (J) | | | |
| CON-RT | 24.6 ± 6.8 | 26.0 ± 9.0 | 0.24 |
| ECC-RT | 21.8 ± 5.4 | 25.8 ± 7.0* | 0.64 |
| Concentric work 90°/s (J) | | | |
| CON-RT | 23.4 ± 6.7 | 23.9 ± 7.8 | 0.08 |
| ECC-RT | 21.5 ± 5.1 | 24.3 ± 4.8* | 0.47 |

**Notes:** CON-RT = concentric resistance training; ECC-RT = eccentric resistance training; PT = peak torque; ES = effect size; *P < 0.05 vs. Pre.

**Table S6.** Training effects on dorsiflexion eccentric strength parameters.

| | Pre | Post | ES |
|---|---|---|---|
| ***Preceded by an isometric contraction*** | | | |
| Eccentric PT 10°/s (Nm) | | | |
| CON-RT | 53.1 ± 14.5 | 55.5 ± 15.3 | 0.18 |
| ECC-RT | 50.8 ± 13.4 | 55.9 ± 12.0* | 0.37 |
| Eccentric PT 45°/s (Nm) | | | |
| CON-RT | 52.3 ± 12.6 | 53.8 ± 16.2 | 0.13 |
| ECC-RT | 50.0 ± 12.0 | 55.4 ± 12.3*# | 0.45 |
| Eccentric PT 90°/s (Nm) | | | |
| CON-RT | 50.3 ± 13.8 | 51.4 ± 16.6 | 0.09 |
| ECC-RT | 48.3 ± 11.4 | 55.0 ± 11.4*# | 0.54 |
| Average net eccentric PT (Nm) | | | |
| CON-RT | 6.9 ± 4.1 | 4.0 ± 4.6* | -0.72 |
| ECC-RT | 6.5 ± 4.2 | 9.3 ± 4.0*# | 0.68 |
| Average net eccentric PT (%) | | | |
| CON-RT | 16.5 ± 11.8 | 7.5 ± 11.0* | -0.85 |
| ECC-RT | 15.4 ± 9.8 | 20.5 ± 8.7*# | 0.48 |
| ***Preceded by a concentric contraction*** | | | |
| Eccentric PT 10°/s (Nm) | | | |
| CON-RT | 45.0 ± 11.5 | 47.0 ± 13.5 | 0.18 |
| ECC-RT | 43.4 ± 10.9 | 49.5 ± 11.4*# | 0.56 |
| Eccentric PT 45°/s (Nm) | | | |
| CON-RT | 46.1 ± 10.6 | 47.2 ± 14.2 | 0.10 |
| ECC-RT | 43.4 ± 9.8 | 49.1 ± 11.1*# | 0.57 |
| Eccentric PT 90°/s (Nm) | | | |
| CON-RT | 45.3 ± 11.1 | 46.8 ± 14.6 | 0.15 |
| ECC-RT | 42.7 ± 9.5 | 49.5 ± 11.1*# | 0.67 |

**Notes:** CON-RT = concentric resistance training; ECC-RT = eccentric resistance training; PT = peak torque; ES = effect size; *P < 0.05 vs. Pre. #P < 0.05 greater effect than the other condition.

**Figure S1.** Changes in fascicle length (FL) and isometric peak torque, with Pearson correlations presented with and without the data for the CON-RT subject indicated with an asterisk. Bold line and confidence intervals are plotted for the regression analysis with all data.

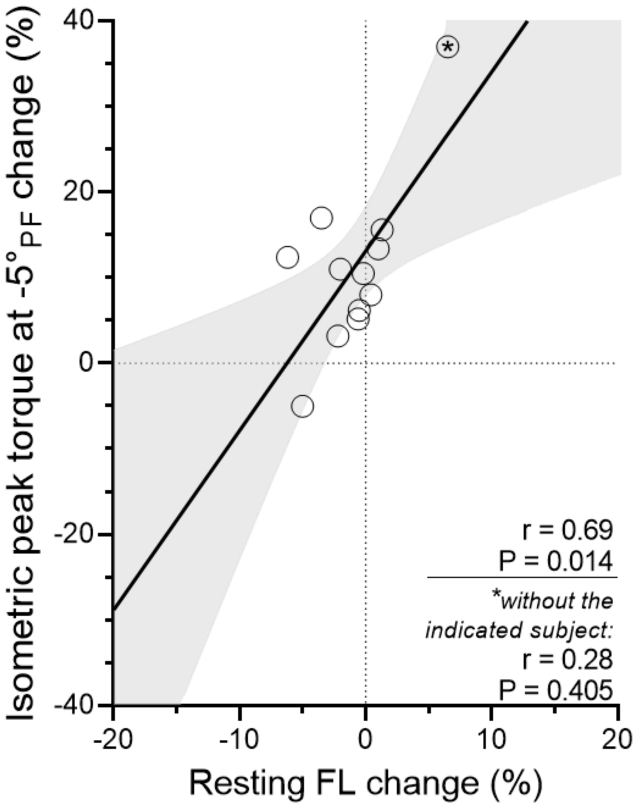

**Figure S2.** Torque–angle–velocity traces (y axes) by time (x axis) obtained from the LabChart software during the tests.

A) Passive:

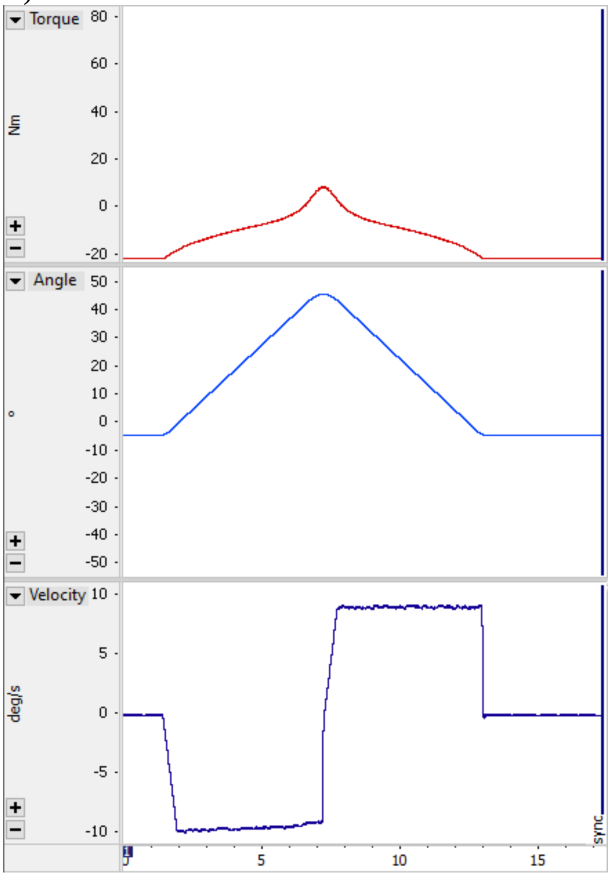


B) Isometric at 20°$_{PF}$:

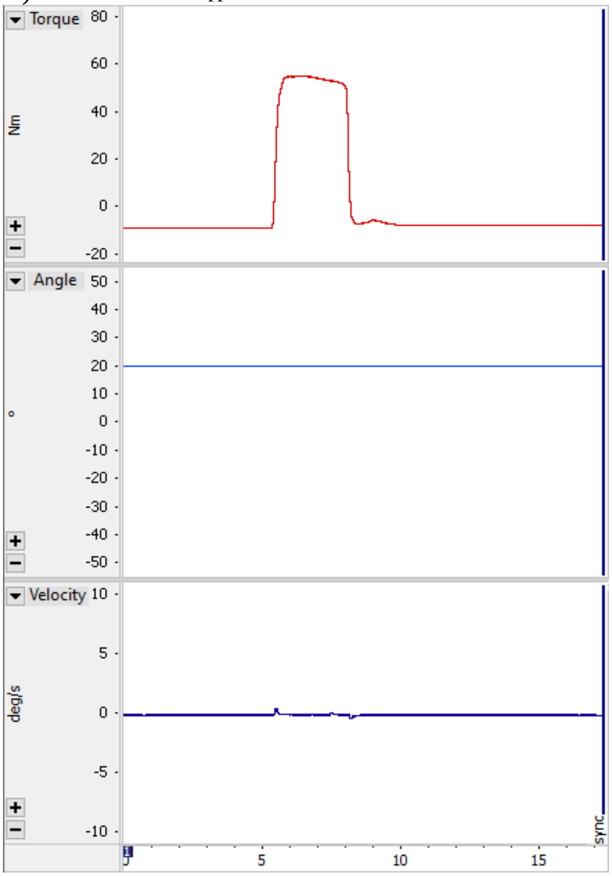


C) Eccentric-concentric pair:

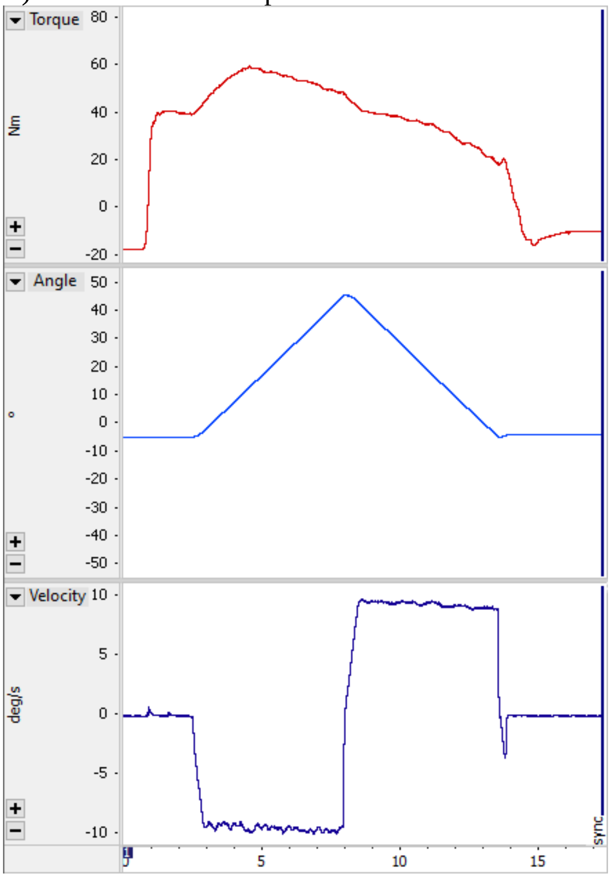


D) Concentric-eccentric pair:

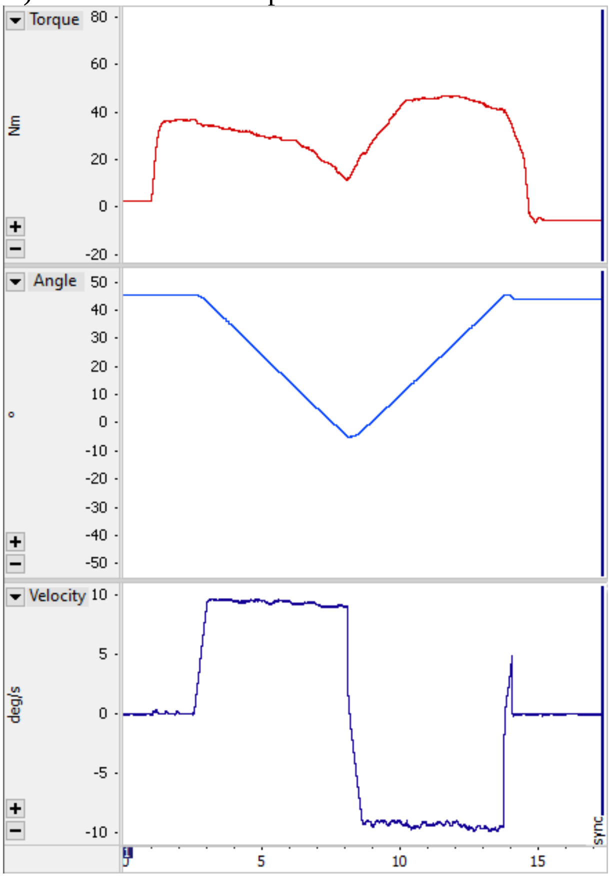
